# Improving estimation of vaccine effectiveness during outbreaks in low-resource settings: A case study of oral cholera vaccination during the 2022-2023 cholera outbreak in Malawi

**DOI:** 10.64898/2026.03.29.26349659

**Authors:** Latif Ndeketa, Daniel Hungerford, Virginia E. Pitzer, Khuzwayo C. Jere, Kondwani C. Jambo, Upendo L. Mseka, Nelson Kumwenda, Chrissy Banda, Matthew Kagoli, Innocent Chibwe, Patrick Musicha, Nigel A. Cunliffe, Neil French, Peter J. Dodd

## Abstract

**Background:** Use of oral cholera vaccine (OCV) is globally recommended as a public health response to cholera outbreaks, alongside water, sanitation and hygiene (WASH) interventions. Estimating vaccine effectiveness during emergencies in low- and middle-income countries is challenging because vaccination campaigns are often implemented over short time frames, while individual-level data are frequently incomplete due to constraints in infrastructure, resources and data systems. There is a need for pragmatic approaches that can generate timely, policy-relevant evidence using routinely collected data.

**Methods:** We analysed routine surveillance data from a large 2022–2023 cholera outbreak in Blantyre District, Malawi. The EpiEstim framework was used to generate estimates of the time-varying reproduction number (Rₜ) from line-listed case data. We modelled changes in 𝑅_𝑡_ as a function of cumulative OCV coverage using a log-linear framework and propagated uncertainty through posterior sampling. Lagged WASH exposure variables were incorporated in the model to generate adjusted vaccine effectiveness estimates and to explore potential interaction effects. Sensitivity analyses assessed robustness to alternative lag structures.

**Findings:** The Blantyre outbreak was characterised by an initial period of low-level transmission followed by a sharp increase in cases from late November 2022, after which transmission declined steadily through April 2023. This decline coincided with the implementation of a reactive OCV campaign. The majority of the cases were among middle-aged men living in urban Blantyre. The unadjusted vaccine-associated reduction in transmission was estimated at 53.52% (95% credible interval (CrI):42.5–64.1%). After adjusting for a 7-day rolling average WASH activity, total vaccine effectiveness increased to 62.1% (95% CrI: 49.3–74.9%). Sensitivity analyses using alternative lag structures for WASH exposure produced comparable adjusted estimates.

**Interpretation:** Implementation of OCV contributed to a substantial reduction in cholera transmission during the outbreak. This study demonstrates a feasible approach for estimating vaccine-attributable impact whilst accounting for public health and social measures, such as WASH interventions. The methods described will be useful in outbreaks where classical observational designs are not possible, providing actionable evidence to policy makers for outbreak response in resource-limited settings.

**Funding:** MRC Discovery Medicine North (DiMeN) Doctoral Training Partnership (UKRI), National Institute for Health and Care Research (NIHR) Global Health Research Group on Gastrointestinal Infections and Wellcome through the core grant to the Malawi-Liverpool-Wellcome Research Programme.

**Research in context:** *Evidence before this study:* Before undertaking this analysis, our team and collaborators generated detailed epidemiological and genomic evidence describing the 2022–2023 cholera outbreak in Blantyre, Malawi. Our previous fine-scale spatial work demonstrated marked heterogeneity in cholera burden across urban Blantyre. We found that transmission concentrated in densely populated informal settlements and areas with poor water and sanitation infrastructure. Our genomic analysis of outbreak isolates showed that the epidemic was driven by a recently introduced seventh-pandemic El Tor *Vibrio cholerae* sub-lineage AFR15 linked to regional and international transmission events, with epidemic expansion occurring during late 2022. These studies characterised where transmission occurred and the evolutionary origin of the outbreak strain. The population-level effect of reactive oral cholera vaccination (OCV) or concurrent and improved water, sanitation and hygiene (WASH) interventions on transmission dynamics during the outbreak was not quantified. In linked work, we conducted a systematic review of post-licensure vaccine impact and effectiveness studies from sub-Saharan Africa (CRD42023436851) to assess the principal study designs and the extent to which they adjusted for concurrent public health and social measures (PHSMs), such as WASH interventions. We searched PubMed, EMBASE, MEDLINE, CINAHL, and Google Scholar for vaccine impact or effectiveness studies conducted in children under five years and screened reference lists of included studies. Across all eligible studies, none measured or adjusted for concurrent PHSMs.

*Added value of this study:* Using surveillance, vaccination, and WASH data collected from Blantyre district by the Malawi Ministry of Health, we estimated the vaccine-associated reduction in transmission during a large reactive OCV campaign. By linking time-varying *R*ₜ estimates to cumulative vaccine coverage within a posterior sampling framework, we provide field-relevant effectiveness estimates from a setting where traditional individual-level designs were not feasible. We further evaluated whether concurrent WASH activity materially altered vaccine effect estimates.

*Implications of all the available evidence:* Taken together with prior spatial and genomic analyses of the same outbreak, our findings provide a more complete picture of cholera control in Malawi. The earlier work clarified transmission geography and pathogen introduction. The present analysis quantifies the population-level impact of reactive vaccination under real-world programme conditions. This integrated evidence base supports ministries of health in interpreting vaccine performance during rapidly evolving outbreaks and in strengthening the routine data systems required for timely evaluation of interventions.

## Introduction

Cholera remains a significant public health concern in low- and-middle-income countries (LMICs) where inadequate water, sanitation, and hygiene (WASH) contribute to recurrent outbreaks(1). Globally, there are an estimated 1.3–4.0 million cases and 143,000 cholera-attributable deaths each year worldwide(2). Cholera is caused by *Vibrio cholerae* (*V. cholerae*), a Gram-negative bacterium with a distinctive curved rod shape. Infection with the bacterium results in a severe and potentially deadly acute diarrhoeal illness(3). *V. cholerae* comprises many serogroups, though epidemic cholera has historically been associated with serogroups O1 and O139. In recent decades, however, nearly all reported outbreaks have been caused by the O1 seventh-pandemic El Tor lineage (7PET) (4,5). Oral cholera vaccines (OCVs) are recommended by the World Health Organization (WHO) for use in addition to other public health and social measures (PHSMs) such as case management and improved WASH(6,7).

Cholera is endemic in Malawi, a low-income African country with recurrent seasonal outbreaks typically occurring during the rainy season between November and May. The number of reported cholera-attributable deaths during seasonal outbreaks in Malawi rarely exceeds 100 deaths nationwide. The largest recorded cholera outbreak occurred between January 2022 and May 2023, resulting in over 58,000 cases and over 1,700 deaths(5,8). This outbreak had a recorded case fatality risk (CFR) of 3.1%, exceeding the CFR observed in neighbouring countries(9). A genomic analysis of the 2022-2023 outbreak revealed the 7PET lineage O1 Ogawa serotype as the primary driver of the outbreaks in Malawi(5). The Ministry of Health of Malawi responded to the cholera outbreak through a combination of reactive vaccination with oral cholera vaccine (OCV) and targeted WASH interventions(10). Vaccination was implemented as part of the national outbreak response strategy. WASH activities were delivered using a case-area targeted intervention (CATI) approach, which focuses on improving hygiene measures around detected cases and within outbreak hotspots(11,12). While OCV deployment aimed to reduce individual susceptibility, WASH measures aimed to limit environmental transmission.

The WHO recommends three prequalified OCVs for use in endemic and outbreak settings(6). All the vaccines contain killed whole *V. cholerae* O1 cells but they differ slightly in formulation and administration. Dukoral™ (Valneva, Sweden) contains inactivated *V. cholerae* O1 cells together with the recombinant B-subunit of cholera toxin. Shanchol™ (Shantha Biotechnics-Sanofi Pasteur, India) and Euvichol™ (EuBiologics, Republic of Korea) are killed whole-cell vaccines, which simplifies field delivery. These two vaccines share the same antigenic composition and have demonstrated comparable safety and immunogenicity across multiple evaluations(3,13). During the 2022–2023 outbreak response in Malawi, Euvichol-Plus®, an improved plastic-vial formulation of Euvichol™, was deployed to facilitate large-scale field delivery. The vaccine has a reported effectiveness ranging from 37% to 80% in various settings including Zambia, India, Bangladesh and Haiti(14–17).

Evaluating the effectiveness of OCV during large-scale outbreaks presents practical and methodological challenges. Rapid vaccination campaigns deployed during an ongoing crisis often rely on routine surveillance that is frequently incomplete or delayed. This lack of timely and high-quality data makes traditional study designs such as case-control or cohort studies difficult to implement, particularly when very few vaccinated cases are observed and when transmission dynamics change faster than recruitment or follow-up periods. Therefore, there is an urgent need for more pragmatic approaches to enable timely measurement of vaccine effectiveness (VE) during emergencies, particularly when other PHSMs, such as WASH, are also introduced.

To address this gap, our study uses a novel approach that leverages the time-varying reproduction number (𝑅_𝑡_ ) to estimate the vaccine-attributable effect of OCV in the presence of concurrent WASH interventions. 𝑅_𝑡_ , which represents the average number of secondary infections generated by a primary case occurring at time *t*, provides evidence for the combined influence of population susceptibility, human behaviour and public health interventions such as vaccination and WASH on cholera transmission, offering a more dynamic and pragmatic way to evaluate VE during emergency responses(18). By adopting this approach using routinely collected data, we aim to pragmatically improve estimation of VE in outbreak settings and provide crucial information for future outbreak response.

## Methods

### Study setting

The analysis focused on the January 2022 to April 2023 cholera outbreak in Blantyre district, Malawi. The district measures 240 km² with a population of approximately 1,400,000 people; healthcare is delivered through a network of private and government health facilities((19,20)). These health facilities also serve as fixed points for routine and reactive immunisation for the expanded programme on immunisation (EPI). Blantyre is divided into five geographically based clusters, each encompassing several health facilities and their catchment areas. This clustering approach is routinely used by district health authorities to plan and monitor public health interventions such as responsive vaccination campaigns and WASH activities.

### Case data

We obtained line-listed anonymised cholera case data from the Blantyre District Health Office (DHO) surveillance system which aggregated case reports from 32 health facilities within Blantyre. Each entry included the date of symptom onset and diagnostic confirmation. A confirmed case was defined as *V. cholerae* confirmed by rapid diagnostic testing of stools plus the clinical case definition of cholera of at least three episodes of watery diarrhoea within the last 24 hours, with or without dehydration or vomiting. A probable cholera case was defined as any clinical case definition plus confirmed contact with a case. Probable case definitions were used when the health system was overwhelmed with high incidence of cases and stool testing was not possible.

### Vaccine data

conducted a reactive oral cholera vaccination campaign from December 2022. Administrative reports indicated coverage of approximately 76% among the eligible population, although the underlying data were not available for verification. For the purposes of this analysis, vaccine coverage was derived from the available time-series data used in the model, which reached a maximum of approximately 24%. Although the vaccine is administered as a two-dose regimen given two weeks apart, constraints in supply meant that most recipients received a single dose. Vaccination counts were reported daily at facility level as doses administered against doses allocated, and dose allocation was estimated through catchment area populations. Coverage was calculated as cumulative doses administered in the district per eligible population over time. The vaccine was offered to all individuals aged one year and older and was administered through a hybrid strategy of routine administration at health facilities and campaigns conducted door-to-door and in populous areas like markets and religious gatherings.

### WASH data

Health authorities adopted the CATI strategy to guide WASH interventions. From December 2022, they disinfected the homes and latrines of confirmed *V. cholerae* cases. Each affected household received two buckets (one for safe water storage and one for hand hygiene) and chlorine-based water treatment agents. Neighbouring households within a defined ring around each case also received the same WASH package. Chlorine tablets were distributed in disease hotspot areas reporting sustained transmission. The WASH intervention teams also conducted environmental surveillance, including water quality testing for faecal contamination and chlorine residual levels. WASH activities were recorded as the number of households the Blantyre DHO teams reached per day.

## Methods of Analysis

### Estimation of time-varying reproduction number (***𝑅_𝑡_*** )

We estimated 𝑅_𝑡_ from daily *V. cholerae* case counts using the *EpiEstim* package in R, which incorporates a user-defined serial interval distribution. We assumed a gamma-distributed serial interval with a mean of 5 days and standard deviation of 8 days(21). The serial interval is defined as the time between symptom onset in successive cases and was obtained from previous Malawi studies(22). EpiEstim returned a posterior distribution for each day representing plausible values of 𝑅_𝑡_ given case incidence and the assumed serial interval.

### Breakpoint identification and analytic window

We used the *strucchange* package in R to identify structural breaks in the weekly time series. This approach was used to identify the earliest significant shift in transmission dynamics, which we interpreted as the transition from the endemic/sporadic phase to the epidemic phase of the outbreak(23). The primary breakpoint was identified on 11 December 2022 (95% CI: 04 December – 18 December). Consequently, all regression analyses for vaccine effectiveness and WASH impact were restricted to the period from 11 December 2022 onwards to focus on the period of sustained transmission.

### Unadjusted vaccine effectiveness

We expect on mechanistic grounds that 𝑅_𝑡_ will vary in proportion to the fraction of the population that is susceptible, i.e.

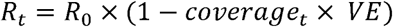

This neglects depletion of susceptibles via natural immunity. Regressing 𝑅_𝑡_ against coverage allows us to extract VE but has the potential to generate negative values of VE for sampled 𝑅_𝑡_ trajectories. To avoid this problem, we assumed that (1 − 𝑐𝑜𝑣𝑒𝑟𝑎𝑔𝑒_𝑡_ × 𝑉𝐸) ≈ 𝑒𝑥𝑝(−𝑐𝑜𝑣𝑒𝑟𝑎𝑔𝑒_𝑡_ × 𝑉𝐸), which is true when coverage times VE is low:

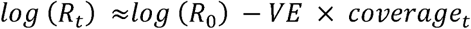

For each of 1,000 𝑅_𝑡_ trajectories, we then fitted a regression of the above form and estimated VE based on the coefficient of the time-varying vaccine coverage. Under this approximation, the slope coefficient provides a direct estimate of vaccine effectiveness, which we express as a percentage. Coverage at time t was defined as the cumulative proportion of the district-level population vaccinated prior to time t (range 0–1). To reflect the delay between vaccination and the development of protective immunity, we applied a seven-day biological lag to vaccine coverage, such that coverage at time t represents the proportion vaccinated at time t−7. The distribution of VE across all posterior samples yielded a median estimate and 95% credible interval (CrI), reflecting uncertainty in both 𝑅_𝑡_ estimation and model fit.

### WASH and vaccine variables

As stated above, WASH activities were recorded as the number of households the Blantyre that DHO teams reached per day. Dates were irregular due to operational deployment patterns. Daily household counts were then aggregated and joined to the cholera time series. Days with no recorded intervention activity were retained as zero events to avoid missing data and to ensure a complete time series. We converted daily WASH counts to households reached per 1,000 population to capture the temporal effect of WASH on transmission. From this, we derived two exposure measures: a 7-day rolling average representing short-term intensity of WASH activity and a cumulative exposure measure capturing the total number of households reached up to each day. We phenomenologically modelled WASH using the short-term measure rather than a cumulative exposure. This reflects the nature of the multiple components involved in WASH activities, where some components are transient while others may persist. A cumulative formulation assumes sustained impact across all components and, in this setting, was highly correlated with vaccine coverage. This reduces the ability of the model to separate their independent effects on transmission. WASH variable was then lagged by seven days because WASH activities improve environmental conditions more gradually(20,21). In addition, we applied a seven-day biological lag to vaccine coverage to reflect the delay between vaccination and the development of protective immunity.

### Adjusted vaccine effectiveness model

To derive a VE estimate adjusted for the concurrent WASH intervention, we modified our regression to represent a phenomenological effect of WASH coverage on 𝑅_𝑡_:

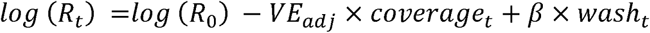

Under the same low-coverage approximation, the coefficient on time-varying coverage provides a direct estimate of VE after adjustment for WASH intensity. This model estimates the vaccine-attributable effect independent of variation in WASH activity, rather than the combined effect of concurrent interventions. The 7-day rolling average WASH variable represents the short-term intensity of households reached over time at district level. However, it does not necessarily represent continuous protection of households already reached. All the variables in the analysis are described in detail in Supplementary Table S1.

### Sensitivity analysis

We assessed the robustness of the results by varying assumptions about the timing of intervention effects. Exposure variables for vaccine coverage and WASH intensity were reconstructed using alternative lag structures of 0 and 14 days. These estimates were compared with the primary specification using a 7-day biological lag. The adjusted vaccine effectiveness model was refitted to all 1,000 𝑅_𝑡_ trajectories under each lag specification. Vaccine effectiveness estimates obtained under the alternative lags were compared with those from the primary 7-day lag model.

### Missing data

Cholera case data with missing RDT results were treated as not tested and included as probable cases. A small number of RDT-negative cases were excluded from the analysis. These decisions did not affect the results. Days without WASH activity were retained as true zero exposure. Vaccine coverage and case reporting were complete at district level for the analytic period. 𝑅_𝑡_ estimation naturally accommodates stochastic noise in case series, and posterior sampling ensures that uncertainty arising from day-to-day variability is propagated into all downstream estimates. No imputation was performed.

## Results

### Demographic characteristics

Between January 2022 and April 2023, a total of 8,013 cholera cases were reported in Blantyre District (Table 1), amounting to 0.6% of the population Cases were concentrated in urban areas and occurred more frequently among males. The age distribution was typical of cholera epidemics, with the highest burden observed among young adults and children and adolescents. Most cases were classified as probable on the basis of epidemiological linkage, with laboratory confirmation limited by testing capacity during the outbreak. The outbreak resulted in 201 deaths, corresponding to a CFR of 2.5%. Vaccination history was rarely documented among reported cases, with fewer than one percent having a recorded history of oral cholera vaccination.

**Table 1:**
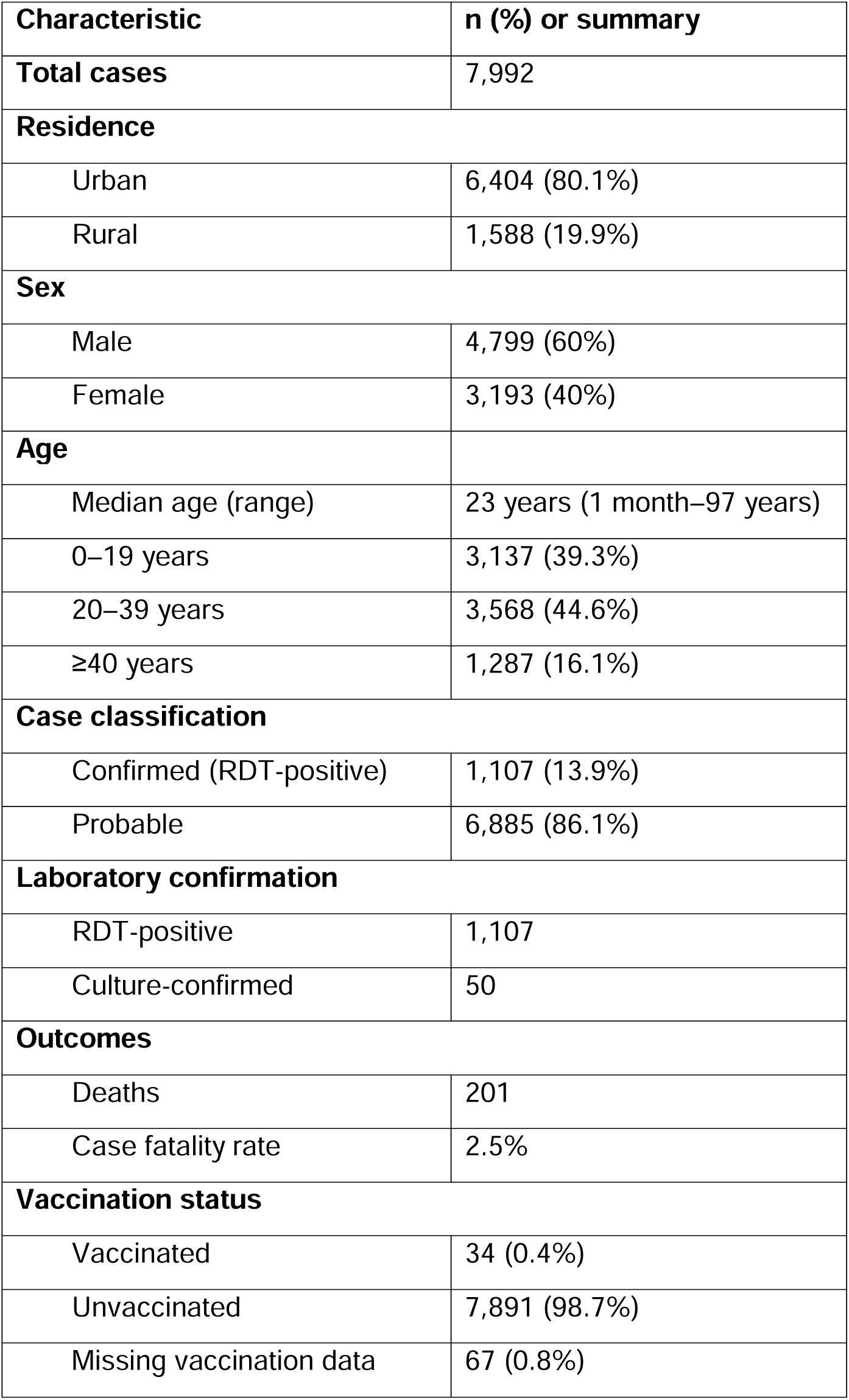
Descriptive characteristics of cholera cases in Blantyre District, Malawi, January 2022 –April 2023.

**Table 2:** Posterior estimates of vaccine coverage and WASH effects on R ₜ.

| Model | $\beta$ (log-HR) median | 95% CrI ($\beta$) | HR median | 95% CrI (HR) | VE (%) median | 95% CrI (VE) |
| --- | --- | --- | --- | --- | --- | --- |
| Vaccine effect only (lagged 7 days) | -0.535 | -0.641 – -0.425 | 0.59 | 0.53 – 0.65 | **53.5** | 42.5 – 64.1 |
| WASH only (lagged 7 days) | -0.009 | -0.011 – -0.007 | 0.991 | 0.989 – 0.993 | — | — |
| Vaccine + WASH (lagged 7 days) | -0.621 | -0.749 – -0.493 | 0.54 | 0.47 – 0.61 | **62.1** | 49.3 – 74.9 |

**Table 3:**
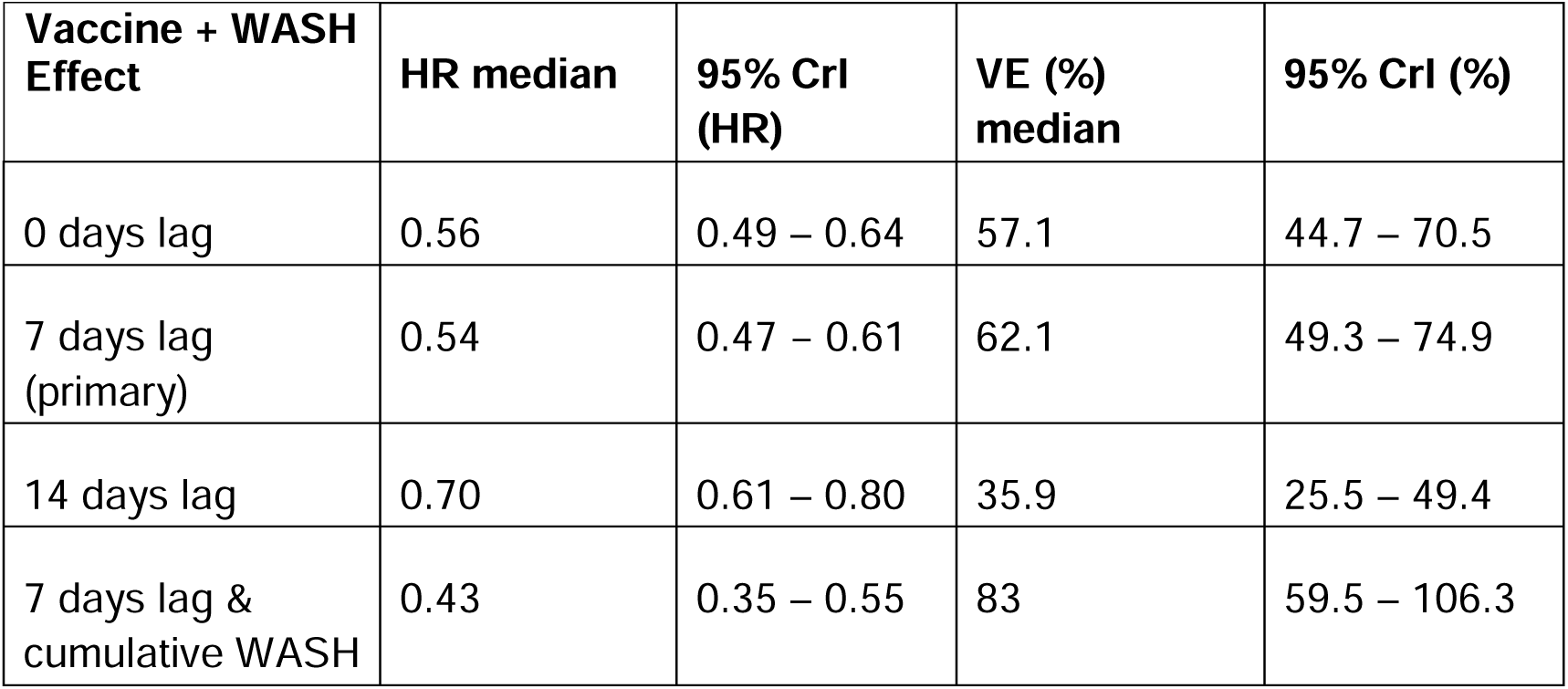
Vaccine effectiveness estimates from the WASH-adjusted model across alternative cumulative WASH lag specifications.

| Vaccine + WASH Effect | HR median | 95% CrI (HR) | VE (%) median | 95% CrI (%) |
| --- | --- | --- | --- | --- |
| 0 days lag | 0.56 | 0.49 – 0.64 | 57.1 | 44.7 – 70.5 |
| 7 days lag (primary) | 0.54 | 0.47 – 0.61 | 62.1 | 49.3 – 74.9 |
| 14 days lag | 0.70 | 0.61 – 0.80 | 35.9 | 25.5 – 49.4 |
| 7 days lag & cumulative WASH | 0.43 | 0.35 – 0.55 | 83 | 59.5 – 106.3 |

Reported cholera cases were geographically heterogeneous across Blantyre, with high burdens observed across multiple health-facility catchments rather than being confined to a single focal area (Figure 1). This pattern indicates that transmission was distributed across several parts of the district rather than being driven by one dominant hotspot.

**Figure 1:**
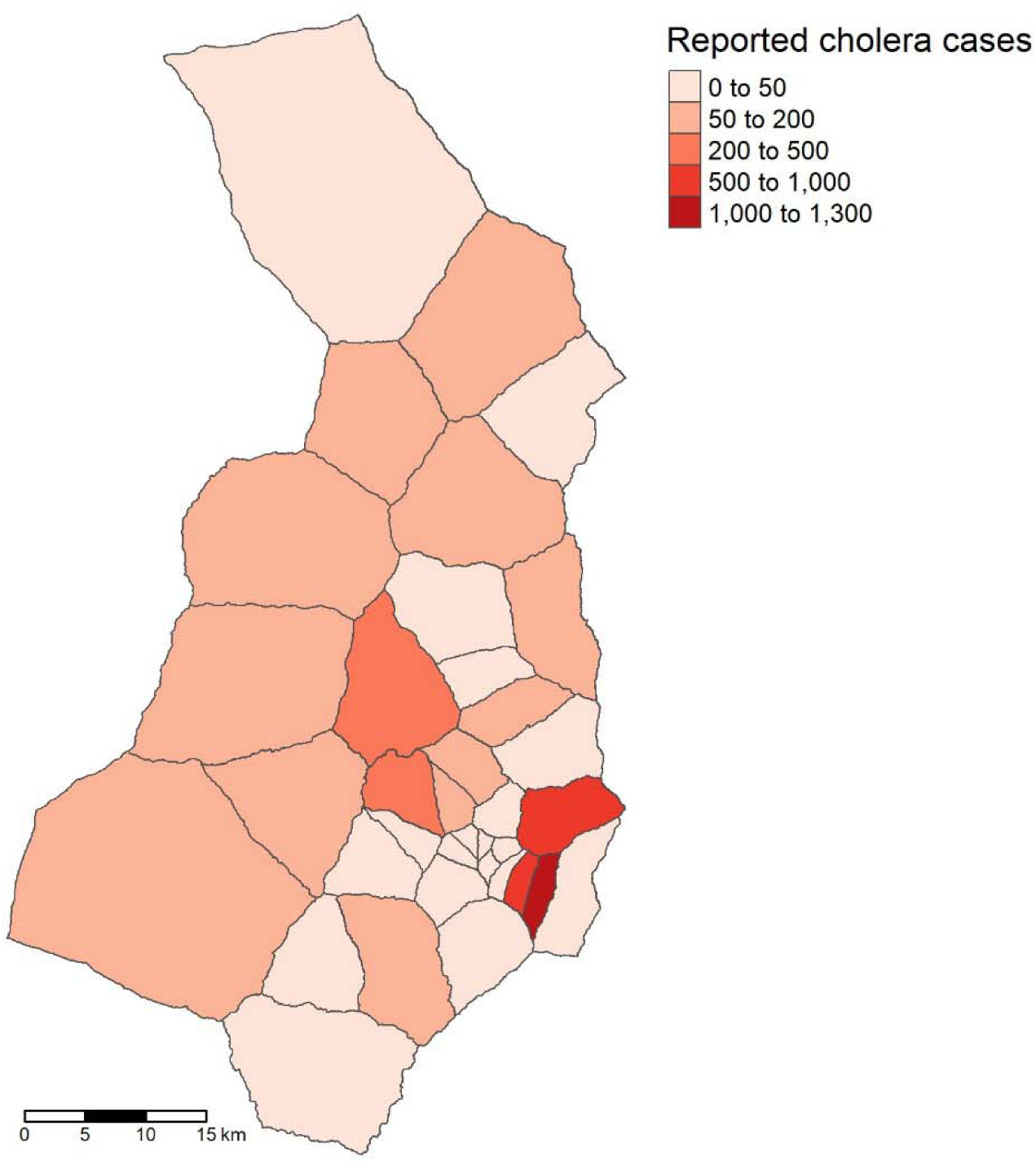
Spatial distribution of reported Blantyre cholera cases by health facility catchment area.

### Outbreak description

As illustrated in Figure 2, the epidemic began with a period of sporadic cases from January 2022 to April 2022. During this phase, the median 𝑅_𝑡_ was 1.74 (95% CrI: 0.12 – 4.30), and 77.6% of weeks were above the epidemic threshold of 𝑅_𝑡_ >1. From May to November 2022, cholera cases occurred consistently, although incidence remained low. The median 𝑅_𝑡_ during this period was 1.03 (95% CrI: 0.68 – 1.50), with 62.1% of weeks above the epidemic threshold. This reflects sustained but low-level transmission. The outbreak reached its peak in December 2022, corresponding to the weeks with the highest number of reported cases (Figure 2).

**Figure 2:**
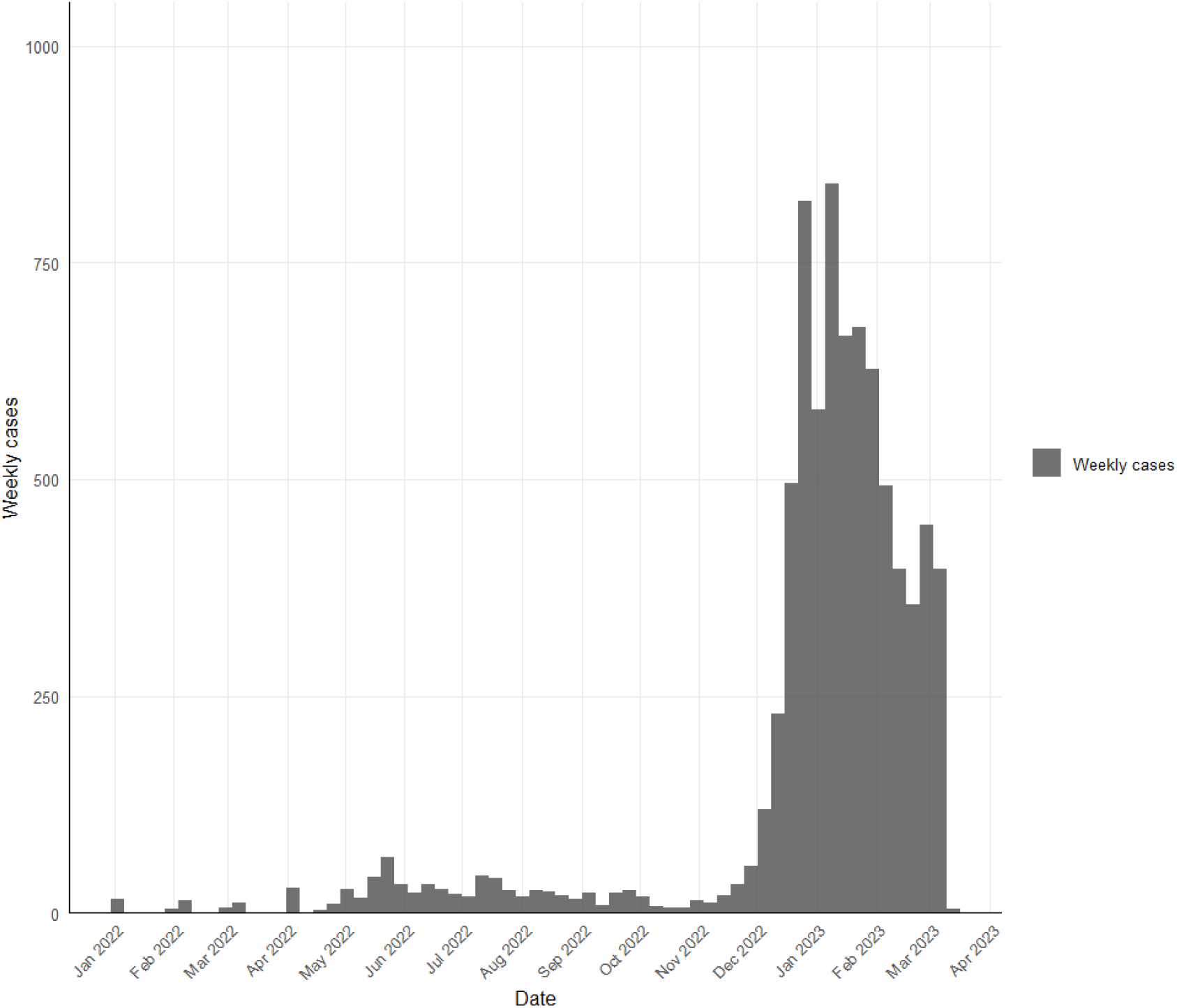
Temporal distribution of cholera cases in Blantyre.

Following this peak, incidence declined gradually until April 2023. During the vaccination period (December 2022–April 2023), the median 𝑅_𝑡_ was 1.0 (95% CrI: 0.92–1.08).

Breakpoint analysis identified 11 December 2022 (95% CI 04 – 18 December) as the point marking the onset of sustained transmission where the weekly number of cases rose from approximately 22.7 to 529.1. Figure 3B illustrates how transmission shifted as vaccination and WASH activities were introduced. Responsive vaccination started in early December 2022 and scaled up rapidly through January as cumulative doses increased across the district (Figure 3C). 𝑅_𝑡_ was already beginning to decline at the outset of this rollout, with the subsequent drop coinciding with sharp gains in vaccine coverage. WASH interventions were initiated later, deployed intermittently from mid-December 2022, and most intensively between January and March 2023 (Figure 3D).

**Figure 3:**
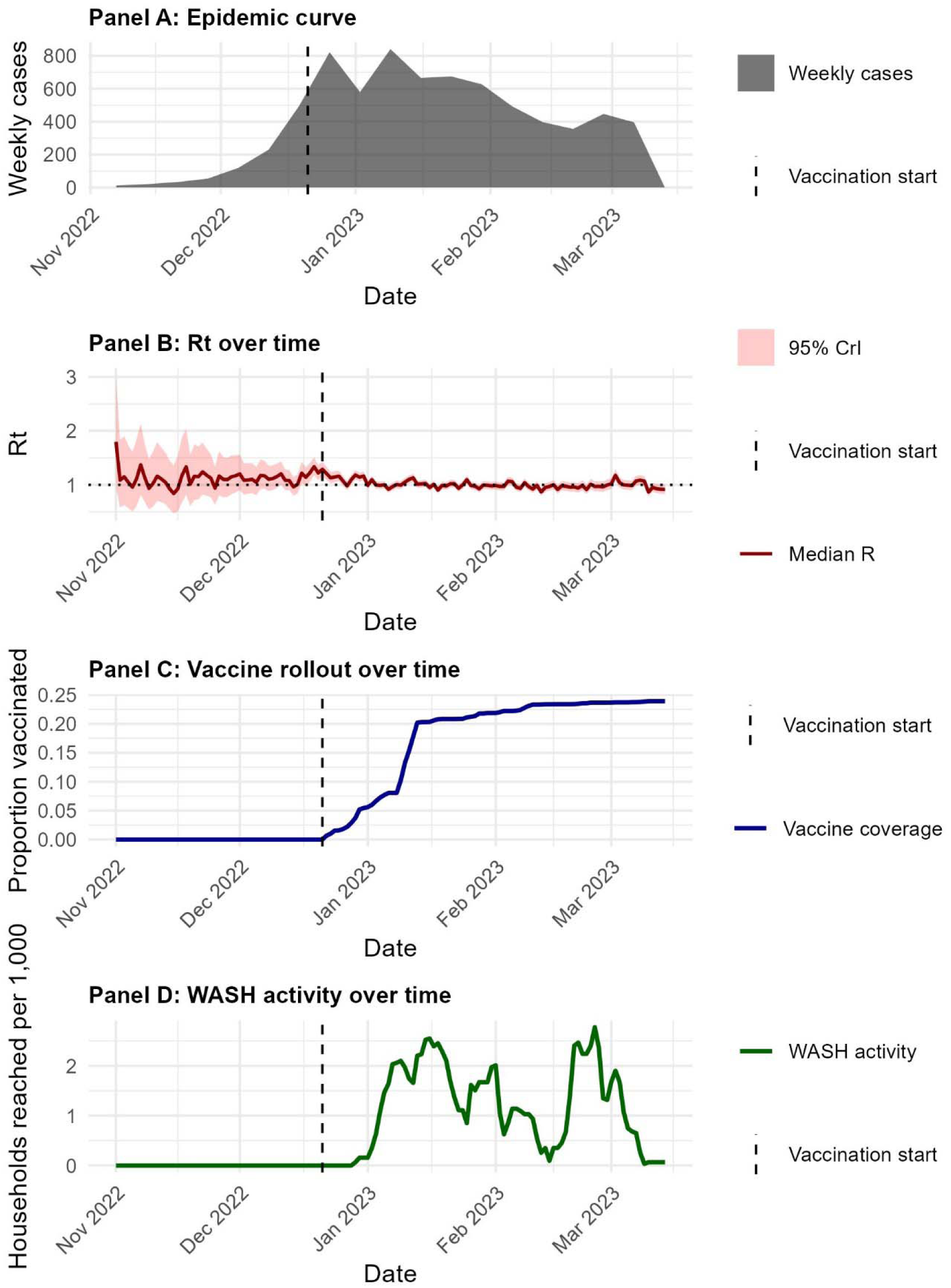
Weekly cholera incidence and time-varying transmission dynamics in Blantyre, alongside the timing of vaccination and WASH interventions from November 2022. Panel A shows weekly reported cases. Panel B shows the median Rₜ with 95% credible intervals. Panel C shows cumulative vaccine coverage. Panel D shows WASH activity intensity, expressed as households reached per 1,000 population. The dashed vertical line marks the start of vaccination

### Vaccine effectiveness

Using 1,000 posterior draws from the 𝑅_𝑡_ distribution, the unadjusted VE was estimated to be 53.5% (95% CrI: 42.5 – 64.1), as shown in Table 1. This represents the estimated reduction in transmission potential associated with 100% vaccine coverage, independent of other interventions. When adjusting for WASH activity, the VE estimates increased to 62.1% (95% CrI: 49.3 – 74.9) which suggested that vaccination retained a strong independent effect even after accounting for concurrent WASH interventions.

In addition, we explored the effect of WASH activities on 𝑅_𝑡_ and found that WASH interventions were associated with a small reduction in transmission intensity. Using the 7-day rolling WASH exposure lagged by seven days, the posterior distribution of the hazard ratio was consistently below unity, with a median estimate of 0.993 (95% CrI: 0.990–0.996).

The sensitivity analyses using alternative lag assumptions and WASH specifications showed variation in estimates. The median VE was 57.1% (95% CrI: 44.7–70.5) with no lag, 62.1% (95% CrI: 49.3–74.9) with a 7-day lag, and 35.9% (95% CrI: 25.5–49.4) with a 14-day lag. Using cumulative WASH exposure in place of the rolling WASH variable produced a higher VE estimate of 83.0% with uncertainty bounds exceeding 100% (95% CrI: 59.5–106.3).

## Discussion

We developed a pragmatic method for estimating vaccine performance during a fast-moving cholera outbreak, with concurrent PHSMs, such as WASH interventions, where traditional epidemiological designs were not feasible. Using routinely collected surveillance data and a framework that links temporal changes in 𝑅_𝑡_ to changes in vaccine coverage, we found in unadjusted analysis that OCV was associated with a reduction of 53.5% in cholera disease transmission. Vaccine effectiveness increased to 62.1% once a lagged 7-day rolling WASH variable was adjusted for in the model. The similarity between unadjusted and adjusted estimates suggests that the decline in transmission was not primarily driven by the reported WASH interventions and that vaccination contributed independently to the epidemic control during the response period.

Our VE estimates align with the broader evidence base for killed OCVs, despite differences in design and setting. A recent systematic review and meta-analysis by Xu *et al*. reported VE of 69% for two doses and 60% for one dose at 12 months following vaccination(24). This review synthesised data from 10 observational studies worldwide (two from Africa), most using case–control or cohort designs to measure individual-level protection against *V. cholerae*. None of the studies in this review reported adjusting for WASH in their analysis. In contrast, our analysis estimates changes in transmission at the population level during an outbreak, using 𝑅_𝑡_ , while accounting for WASH activity(25). This reflects a total vaccine effect, incorporating changes in susceptibility, infectiousness, and indirect protection, rather than individual-level protection against disease measured in case–control or cohort studies.

Our findings are also consistent with evidence from sub-Saharan Africa. A matched case-control study in the Democratic Republic of Congo, evaluating a single dose of Euvichol-Plus, reported a VE of 52.7% (95% CI 31.4–67.4) 12–17 months after vaccination(26). A similar design for Shanchol vaccine in Guinea reported a VE of 86.6% (95% CI 56.7 – 95.8%)(27). These estimates fall withing the range observed here.

Our study uses a similar methodology to a study by Hulland and colleagues (2024) who used EpiEstim and the VaxEstim extension to estimate single-dose OCV effectiveness in Haiti and Cameroon(28). However, there are important distinctions to be noted. Their analysis did not have access to full surveillance line-listing and instead reconstructed weekly case counts by digitising epidemic curves from situation reports and outbreak bulletins. Their estimates suggested OCV1 effectiveness of approximately 75.3% in Haiti and 54.9% in Cameroon without adjusting for other interventions such as WASH activities(28). In contrast, our analysis was at the district-level rather than national and adjusted for WASH activity. In addition, we applied a log-linear regression of Rₜ on cumulative coverage and WASH rather than imposing an assumed fixed relationship between Rₜ, coverage and WASH.

A key contribution of this study lies in the method itself. Case-control and cohort designs were not feasible in this outbreak because only 34 vaccinated cases were identified, resources were unavailable for establishing a control population and a prospective cohort design was incompatible with the duration of the epidemic wave. Time-series approaches requiring extended pre- and post-intervention periods were also unsuitable because there was no clear post-vaccination period in our dataset. The posterior sampling framework instead draws information directly from the epidemic curve, propagates uncertainty from 𝑅_𝑡_ estimation, and exploits temporal variation in vaccination coverage to generate population-level estimates of VE(21). This approach does not replace classical designs but offers a feasible alternative for Ministries of Health during emergencies, particularly in low-resource settings where rapid evaluation is crucial and field data collection is constrained.

Interpretation of our findings must also consider the role of naturally acquired immunity. Infection-induced protection develops after symptomatic and asymptomatic *V. cholerae* infection and lasts for at least 3 years(29). In this outbreak, however, the scope for natural immunity to drive large reductions in transmission at district level was probably limited. The 7,992 reported cases represent well under 1% of Blantyre’s population of about 1.4 million, and much of the burden occurred during the peak rather than during the earlier sporadic and low-incidence phases. Even allowing for underreporting and asymptomatic infections, the proportion infected is unlikely to have reached levels where susceptible depletion alone would largely be responsible for the decline in 𝑅_𝑡_ .

The spatial distribution of cases also limits the extent to which the decline can reasonably be attributed to infection-derived natural immunity. Transmission was spread across several clusters rather than being concentrated within a cluster, which makes it unlikely that any one pool of susceptible individuals could have been depleted quickly enough to drive a coordinated fall in 𝑅_𝑡_ at the district level. Infection-derived immunity may still have mattered in particular high-risk areas, yet such effects would have been localised and, on their own, are unlikely to account for the sustained decline in Rₜ observed across Blantyre during the response period. Exploring these local immunity dynamics in more detail will require mechanistic modelling of susceptible depletion which was beyond the scope of this analysis.

The effect of WASH intervention alone on 𝑅_𝑡_ was small with posterior coefficients close to the null. This result should be interpreted with caution as 𝑅_𝑡_ is a distal outcome. The WASH dataset captured only the administrative coverage of CATI activities but did not capture behavioural uptake, sustained use, or changes in environmental contamination at the household level. Furthermore, WASH interventions were delivered intermittently, in a short period and at a stage when transmission was already declining which therefore biases the effect towards the null. Under such conditions, even an intervention with genuine biological and environmental impact may leave only a weak statistical signal when evaluated against 𝑅_𝑡_ at district level.

The effect of WASH in our analysis aligns with the wider literature, which has consistently noted the difficulty of demonstrating population-level effects of WASH interventions on cholera incidence or transmission in emergency settings(30,31). This is particularly evident when evaluations depend on aggregate epidemiological outcomes rather than indicators such as water quality or behavioural adherence. Water quality at point-of-use is the WASH intervention’s target that may help to reduce risk of transmission and disease burden during outbreaks, albeit with moderate quality evidence(25).

Our study was not without limitations. Vaccine coverage was measured ecologically at district level using doses delivered, meaning the effect we estimate reflects population-level impact of 1 or more doses rather than individual protection. In addition, WASH data captured operational activity but not adherence or real-world uptake of behaviours promoted during CATI implementation which may underestimate its impact.

Despite these constraints, the study has several strengths. It offers a method that can be deployed in real time using data routinely available to health authorities without the need for recruitment or individual-level linkage. During an outbreak, this approach could support programme monitoring by providing early indications of whether vaccination campaigns and complementary interventions are associated with reductions in transmission. Such information may assist response teams in assessing the timing and intensity of control measures and in guiding decisions on whether additional vaccination rounds or intensified WASH activities are required. The approach incorporates uncertainty rather than masking it and produces interpretable estimates in settings where evidence is urgently needed to guide programme decisions. The inclusion of WASH activity, even at an ecological level, provides a more realistic estimate of vaccine-attributable impact than models that ignore concurrent interventions.

## Conclusion

This study demonstrates a feasible approach for estimating vaccine-attributable impact whilst accounting for public health and social measures, such as WASH. Using these methods, we show that OCV contributed to a substantial reduction in cholera transmission during the outbreak. More broadly, the work demonstrates that policy makers in LMICs can generate meaningful field VE estimates in resource-constrained settings during outbreaks where traditional observational designs are not feasible and timely evidence is nonetheless required to guide outbreak response.

## Data Availability

All data produced in the present study are available upon reasonable request to the authors

## Funding

This work was supported by the UK National Institute for Health and Care Research (NIHR) Global Health Research Group on Gastrointestinal Infections at the University of Liverpool using UK aid from the UK Government to support global health research (NIHR133066). This work was supported by the Malawi-Liverpool-Wellcome Programme Core Grant from Wellcome (grant number 206545/Z/17/Z). Latif Ndeketa is supported by a studentship from the MRC Discovery Medicine North (DiMeN) Doctoral Training Partnership (MR/W006944/1). Nigel Cunliffe is a NIHR Senior Investigator (NIHR203756) and Khuzwayo Jere is an NIHR Global Health Professor (NIHR306394). Peter Dodd acknowledges research funding through The UK-South East Asia Vaccine Manufacturing Research Hub. This research is funded by the Department of Health and Social Care using UK International Development funding and is managed by the EPSRC. The views expressed in this publication are those of the authors and not necessarily those of the Department of Health and Social Care. Nigel Cunliffe and Daniel Hungerford are also affiliated with the NIHR Health Protection Research Unit in Gastrointestinal Infections at the University of Liverpool, a partnership with the UK Health Security Agency in collaboration with the University of Warwick. The funders had no role in study design, data collection and analysis, decision to publish, or preparation of the manuscript. The content is solely the responsibility of the authors and does not necessarily represent the official views of the National Institutes of Health, the NIHR, the Department of Health and Social Care, the UK government or the UK Health Security Agency.

## Conflicts of interest

D.H. and N.F are currently receiving grant support from Seqirus UK Ltd for the evaluation of influenza vaccines and grant support and personal consultancy fees from Merck & Co (Kenilworth, New Jersey, USA) for rotavirus strain surveillance; D.H. received honoraria for a presentation at a Merck Sharp & Dohme (UK) Limited symposium on vaccines; and D.H. has received research grant support from Sanofi Pasteur, and Merck & Co for rotavirus strain surveillance. K.C.J. has received research grant support from GlaxoSmithKline Biologicals for work on rotavirus vaccines. All other authors declare no competing interests.

## Contributions

L.N, D.H, N.F, N.A.C and P.D conceived of the study and secured the funding. L.N. performed the analysis, wrote the manuscript. N.F, V.E.P, D.H, P.D assisted with the analysis and interpretation of results and edited the manuscript. K.C.J. M.K., C.B, P.M, U.M, N.K and K.C.J oversaw the data collection, assisted with the interpretation of results and edited the manuscript. All authors assisted with the interpretation of results and edited the manuscript.

## Notes

### Competing Interest Statement

The authors have declared no competing interest.

### Author Declarations

This study used routinely collected surveillance data from the Malawi Ministry of Health. The data were anonymised prior to analysis and were provided for secondary use. Formal ethical approval was not required under local guidelines for analysis of de-identified routine public health surveillance data.

## References

1. World Health Organization. Cholera vaccines: WHO position paper – August 2017. Relev Epidemiol Hebd. 2017;92(34):477–98.

2. Ganesan D, Gupta S Sen, Legros D. Cholera surveillance and estimation of burden of cholera. Vaccine. 2020 Feb 29;38:A13–7.

3. Clemens JD, Shin S, Sah BK, Sack DA. Cholera vaccines [Internet]. Sixth Edit. Vaccines: Sixth Edition. Elsevier Inc.; 2012. 141–152 p. Available from: 10.1016/B978-1-4557-0090-5.00023-9

4. Nelson EJ, Harris JB, Morris JG, Calderwood SB, Camilli A. Cholera transmission: the host, pathogen and bacteriophage dynamic. Nat Rev Microbiol [Internet]. 2009 [cited 2025 Jan 29];7(10):10.1038/nrmicro2204. Available from: https://pmc.ncbi.nlm.nih.gov/articles/PMC3842031/

5. Chaguza C, Chibwe I, Chaima D, Musicha P, Ndeketa L, Kasambara W, et al. Genomic insights into the 2022–2023Vibrio cholerae outbreak in Malawi. Nat Commun [Internet]. 2024 Dec 1 [cited 2025 Jan 29];15(1):6291. Available from: https://pmc.ncbi.nlm.nih.gov/articles/PMC11282309/

6. World Health Organization. Immunization, Vaccines and Biologicals [Internet]. 2017 [cited 2025 Dec 17]. Available from: https://www.who.int/teams/immunization-vaccines-and-biologicals/diseases/cholera

7. World Health Organization. Cholera [Internet]. 2024 [cited 2025 Dec 17]. Available from: https://www.who.int/news-room/fact-sheets/detail/cholera

8. Miggo M, Harawa G, Kangwerema A, Knovicks S, Mfune C, Safari J, et al. Fight against cholera outbreak, efforts and challenges in Malawi. Heal Sci Reports [Internet]. 2023 Oct 1 [cited 2025 Jan 29];6(10):e1594. Available from: https://pmc.ncbi.nlm.nih.gov/articles/PMC10551271/

9. World Health Organization. Cholera in the WHO African Region | WHO | Regional Office for Africa [Internet]. 2024 [cited 2026 Feb 17]. Available from: https://www.afro.who.int/health-topics/disease-outbreaks/cholera-who-african-region?page=0

10. UNICEF. Malawi Cholera Response Flash Update - 20 to 26 February 2023 | UNICEF Malawi [Internet]. 2023 [cited 2025 Dec 17]. Available from: https://www.unicef.org/malawi/reports/malawi-cholera-response-flash-update-20-26-february-2023

11. Dunoyer J, Ratnayake R, Moore S, Bulit G, Beaulieu S, Valingot C, et al. Optimizing the implementation of case-area targeted interventions during cholera outbreaks with context-specific delivery mechanisms. PLoS Negl Trop Dis [Internet]. 2025 Sep 1 [cited 2025 Dec 17];19(9):e0013534. Available from: https://journals.plos.org/plosntds/article?id=10.1371/journal.pntd.0013534

12. Finger F, Bertuzzo E, Luquero FJ, Naibei N, Touré B, Allan M, et al. The potential impact of case-area targeted interventions in response to cholera outbreaks: A modeling study. PLoS Med [Internet]. 2018 Feb 1 [cited 2025 Dec 17];15(2):e1002509. Available from: https://pmc.ncbi.nlm.nih.gov/articles/PMC5828347/

13. Ryan ET, Leung DT, Jensen O, Weil AA, Bhuiyan TR, Khan AI, et al. Systemic, Mucosal, and Memory Immune Responses following Cholera. Trop Med Infect Dis [Internet]. 2021 Dec 1 [cited 2025 Dec 17];6(4):192. Available from: https://pmc.ncbi.nlm.nih.gov/articles/PMC8628923/

14. Ivers LC, Hilaire IJ, Teng JE, Almazor CP, Jerome JG, Ternier R, et al. Effectiveness of reactive oral cholera vaccination in rural Haiti: a case-control study. Lancet Glob Heal [Internet]. 2015 Mar 1 [cited 2026 Jan 15];3(3):e162. Available from: https://pmc.ncbi.nlm.nih.gov/articles/PMC4384694/

15. Qadri F, Ali M, Chowdhury F, Khan AI, Saha A, Khan IA, et al. Feasibility and effectiveness of oral cholera vaccine in an urban endemic setting in Bangladesh: A cluster randomised open-label trial. Lancet [Internet]. 2015 Oct 3 [cited 2026 Jan 15];386(10001):1362–71. Available from: https://www.thelancet.com/action/showFullText?pii=S0140673615611400

16. Sialubanje C, Kapina M, Chewe O, Matapo BB, Ngomah AM, Gianetti B, et al. Effectiveness of two doses of Euvichol-plus oral cholera vaccine in response to the 2017/2018 outbreak: a matched case–control study in Lusaka, Zambia. BMJ Open [Internet]. 2022 Nov 11 [cited 2026 Jan 15];12(11):e066945. Available from: https://pmc.ncbi.nlm.nih.gov/articles/PMC9660660/

17. Wierzba TF, Kar SK, Mogasale V V., Kerketta AS, You YA, Baral P, et al. Effectiveness of an oral cholera vaccine campaign to prevent clinically-significant cholera in Odisha State, India. Vaccine [Internet]. 2015 May 15 [cited 2026 Jan 15];33(21):2463–9. Available from: https://www.sciencedirect.com/science/article/pii/S0264410X15003928?via%3Dihub

18. Nash RK, Nouvellet P, Cori A. Real-time estimation of the epidemic reproduction number: Scoping review of the applications and challenges. PLOS Digit Heal [Internet]. 2022 Jun 1 [cited 2025 Dec 17];1(6):e0000052. Available from: https://pmc.ncbi.nlm.nih.gov/articles/PMC9931334/

19. Malawi National Statistics Office. Population Projections 2018 - 2050 [Internet]. 2018 [cited 2025 Dec 18]. Available from: https://www.nsomalawi.mw/census/2018

20. Severe Malaria Observatory. Malawi health system | smo [Internet]. 2020 [cited 2025 Dec 18]. Available from: https://www.severemalaria.org/countries/malawi/malawi-health-system

21. Cori A, Ferguson NM, Fraser C, Cauchemez S. A New Framework and Software to Estimate Time-Varying Reproduction Numbers During Epidemics. Am J Epidemiol [Internet]. 2013 Nov 1 [cited 2025 Feb 4];178(9):1505–12. Available from: 10.1093/aje/kwt133

22. Chabuka L, Choga WT, Mavian CN, Moir M, Tegally H, Wilkinson E, et al. Genomic epidemiology of the cholera outbreak in Malawi 2022-2023. medRxiv [Internet]. 2023;2(2):2023.08.22.23294324. Available from: https://www.medrxiv.org/content/10.1101/2023.08.22.23294324v1 https://www.medrxiv.org/content/10.1101/2023.08.22.23294324v1.abstract

23. Bai J, Perron P. Estimating and Testing Linear Models with Multiple Structural Changes. Econometrica. 1998 Jan;66(1):47.

24. Xu H, Tiffany A, Luquero FJ, Kanungo S, Bwire G, Qadri F, et al. Protection from killed whole-cell cholera vaccines: a systematic review and meta-analysis. Lancet Glob Heal [Internet]. 2025 Jul 1 [cited 2025 Dec 16];13(7):e1203. Available from: https://pmc.ncbi.nlm.nih.gov/articles/PMC12208782/

25. Elizabeth Halloran M, Struchiner CJ, Longini IM. Study Designs for Evaluating Different Efficacy and Effectiveness Aspects of Vaccines. Am J Epidemiol [Internet]. 1997 Nov 15 [cited 2026 Feb 27];146(10):789–803. Available from: 10.1093/oxfordjournals.aje.a009196

26. Malembaka EB, Bugeme PM, Hutchins C, Xu H, Hulse JD, Demby MN, et al. Effectiveness of one dose of killed oral cholera vaccine in an endemic community in the Democratic Republic of the Congo: a matched case-control study. Lancet Infect Dis. 2024;24(5):514–22.

27. Luquero FJ, Grout L, Ciglenecki I, Sakoba K, Traore B, Heile M, et al. Use of Vibrio cholerae vaccine in an outbreak in Guinea. N Engl J Med [Internet]. 2014 May 29 [cited 2026 Mar 8];370(22):2111–20. Available from: https://pubmed.ncbi.nlm.nih.gov/24869721/

28. Hulland EN, Charpignon M-L, El Hayek GY, Zhao L, Desai AN, Majumder MS. Estimating time-varying cholera transmission and oral cholera vaccine effectiveness in Haiti and Cameroon, 2021-2023. medRxiv [Internet]. 2024 Jun 13 [cited 2025 Dec 16];2024.06.12.24308792. Available from: https://pmc.ncbi.nlm.nih.gov/articles/PMC11343247/

29. Leung T, Matrajt L. Protection afforded by previous Vibrio cholerae infection against subsequent disease and infection: A review. PLoS Negl Trop Dis [Internet]. 2021 May 1 [cited 2025 Dec 16];15(5):e0009383. Available from: https://journals.plos.org/plosntds/article?id=10.1371/journal.pntd.0009383

30. Taylor DL, Kahawita TM, Cairncross S, Ensink JHJ. The Impact of Water, Sanitation and Hygiene Interventions to Control Cholera: A Systematic Review. PLoS One [Internet]. 2015 Aug 18 [cited 2025 Dec 17];10(8):e0135676. Available from: https://pmc.ncbi.nlm.nih.gov/articles/PMC4540465/

31. Brown J, Cavill S, Cumming O, Jeandron A. Water, sanitation, and hygiene in emergencies: summary review and recommendations for further research. Waterlines. 2012 Jan 1;31(1):11–29.

