## Supplementaty tables for "Improving estimation of vaccine effectiveness during outbreaks in low-resource settings: A case study of oral cholera vaccination during the 2022-2023 cholera outbreak in Malawi"

**Supplementary Table S1**. Definition of variables used in the Rt–vaccine effectiveness analysis

| **Variable** | **Description** | **Source** | **Role in analysis** |
| --- | --- | --- | --- |
| weekly_cases | Weekly number of suspected and confirmed cholera cases | Blantyre District Health Office (BDHO) line list | Epidemic description |
| Median_R (Rt) | Median time-varying reproduction number estimated using EpiEstim | Derived from case time series | Primary outcome |
| R_lower | Lower bound of the 95% credible interval for Rt | EpiEstim output | Uncertainty quantification |
| R_upper | Upper bound of the 95% credible interval for Rt | EpiEstim output | Uncertainty quantification |
| coverage | Cumulative proportion of district population receiving OCV | BDHO vaccination reports | Main exposure. Proportion (0-1) |
| hh_reached | Number of households reached by WASH teams per day | BDHO WASH activity reports | Raw WASH input |
| hh_reached_k | Daily households reached scaled per 1,000 households | Derived from WASH data | Standardised WASH metric |
| wash_roll7_k | Seven-day rolling sum of households reached per 1,000 | Derived | Short-term WASH intensity |
| wash_roll7_k_lag7 | Seven-day rolling WASH exposure lagged by seven days | Derived | Lagged WASH exposure |
| wash_cum_k | Cumulative households reached per 1,000 households | Derived | Total WASH reach |
| wash_cum_k_lag7 | Cumulative WASH exposure lagged by seven days | Derived | Primary WASH covariate in adjusted model |
| Rt_sample | Simulated Rt draw for each time point | Posterior sampling | Uncertainty propagation |
